# MEASUREMENT EFFICIENCY OF AN INSTRUMENTED MOUTHGUARD UNDER A LARGE RANGE OF HEAD ACCELERATIONS AND THE EFFECTS OF FILTERING

**DOI:** 10.1101/2022.10.20.22281312

**Authors:** Christopher M. Jones, Kieran Austin, Simon Augustus, Kieran Jai Nicholas, Xiancheng Yu, Claire Baker, Mike Loosemore, Mazdak Ghajari

## Abstract

Instrumented mouthguards (iMG) measure head kinematics in sport, but their measurements have not been validated at high levels of accelerations observed in those sports like rugby. In addition, the effects of filter cut-off frequency on the measured kinematics are still unknown. To address these questions, a drop testing helmeted head-form of an anthropometric testing device (ATD) was used to produce a range of accelerations and accurately control them. Peak linear acceleration (PLA), rotational velocity (PRV), rotational acceleration (PRA) and maximum principal strain (MPS) values were computed. The influence of filter cut-off frequency on peak kinematics was also calculated. Comparison of the peak values across ATD and iMG indicated high levels of agreement, with a total concordance correlation coefficient of 0.97 and intraclass correlation coefficients of 0.990 for PLA, 0.970 for PRV, 0.945 for PRA, and 0.970 for MPS. Cut-off frequencies of 100-300Hz did not significantly attenuate peak kinematics, but frequencies lower than 100Hz did. This is the first study to test an iMG under impact conditions seen in sport. The method presented can be used for in-lab validation of iMGs under head accelerations seen in sport. Furthermore, these results can contribute towards defining standards for filtering iMG data.

## INTRODUCTION

Contact and collision sports such as boxing and rugby union expose athletes to mild traumatic brain injuries as a result of head impacts ^19^. Recent evidence has started to relate head impact exposure (HIE) with negative long-term effects such as the neurodegenerative disease chronic traumatic encephalopathy (CTE) ^21,37,41,43,49^. However, there is still a lack of real-world traumatic brain injury (TBI) data paired with head impact kinematics during contact sports. Such data will allow for exploring the links between HIE and TBI.

The growth and advancement of instrumented mouthguards (iMG) has enabled the measurement of head impact kinematics of contact sport athletes in training and competition^10,28,30,50^. There are a number of iMGs that have been used to further understand the HIE in sport^6,10,50,51^. Most mouthguards reported in the literature achieve values that are in close agreement to the anthropometric testing device (ATD) reference systems^10,30,31,35,48^. However, a recent comparison study found that on-field kinematics for different iMGs were materially different, in addition to a number of iMGs reportedly exceeding the laboratory testing ranges ^30^. Though it was beyond the scope of the study to outline why there were differences, a number of contributing factors were suggested such as the trigger thresholds, mandible action upon impact, adherence to teeth, vocalisation and data processing techniques. Stitt et al.^48^ completed a laboratory validation which tested an iMG system over a 20 - 80 *g* magnitude with impact durations ranging from 15-60 *ms*. While authors found high levels of agreement with a Lin’s concordance correlation coefficient (CCC) of 0.997, on field head impact events last for <10 *ms* ^40,50^ and can exceed 100 *g* ^30,40^. All individual validation testing to date has not exceeded 100 *g* ^6,10,24,35,48^ while impacts with higher linear accelerations occur in sporting and are often related to an increased risk of injury ^9,25,27,39^. For instance, dynamic changes in blood brain barrier regulation have been reported in professional mixed martial arts fighters who sustained impacts over 100 *g*^39^, and impacts of up to 146 *g* have been reported within concussed American Football players ^9^. Whilst debate still continues on the efficacy or accuracy of biomechanical ‘thresholds’ for mild TBI, it is clear that impacts of high magnitude do occur. If these impacts are to be utilised as inputs into brain injury criteria and models, it is crucial that iMG measurements are valid and reliable within this range.

Previous work has shown that large differences can occur to the peak linear acceleration (PLA) and peak rotational acceleration data (PRA) when different filters and filter cut off frequencies are utilised ^45^. Specifically, shorter duration and high magnitude head accelerations are most affected by these varying cut off frequencies^45^, while these accelerations typically exceed lab testing ranges ^24,30,48^. These accelerations carry a greater likelihood of inducing traumatic brain injury. Hence, if data processing procedures cause an underestimation of their magnitudes, the confidence with which injurious impacts can be identified is reduced, and less confidence can be placed on brain computational models that utilise these accelerations as inputs. While most commercial iMG systems utilise proprietary data treatment techniques, these are often ‘black box’ processes that can alter output peak impact kinematics unbeknownst to users, and ultimately could lead to the underestimation of kinematics and brain modelling based metrics of TBI.

One aim of valid and reliable iMG systems is to provide input data for accurate brain modelling. Previous work has found that the rotational kinematics significantly affected strain predicted by a range of brain models whilst the linear acceleration did not ^8^. It is therefore essential to test instrumented mouthguard devices under conditions that emphasise the rotational kinematics produced by head impacts. Comparison testing of iMGs to date has predominately focused on linear impact kinematics via pendulums and pneumatic impactors ^6,30,32,48^. To the authors knowledge, the influence of iMG on brain strain estimations when compared to the ATD gold standard has been assessed in a single study, whereby iMGs showed a mean relative error in brain strain of 7.5% - 8.9% ^35^, and one of the measured iMGs did not have a sufficient time sampling window for input into the simulation model. While some iMGs may simply be utilised for on-field evaluation of number and intensity of impacts, their efficacy in predicting brain injury measures should be included in holistic system evaluations, as the wider long-term view of researchers and sporting governing bodies and organisations is to understand the effects of head impact exposure on the brain.

Being able to validate instrumented mouthguards in laboratory conditions that are more representative of those reported on field (i.e. in terms of duration and magnitude), and those that test rotational kinematics to a greater degree should be a crucial step in the development of any iMG that plans for clinical implementation. In addition, not only should impact acceleration be compared but also brain strain predictions to understand the effects of real-world head impact kinematics on the brain during contact sports. Furthermore, how the data collected by such systems is processed should also be carefully and openly evaluated. Therefore, the current study had two main aims: i) to compare the iMG against a gold standard measure of impact kinematics and brain simulation metrics under conditions that are more representative of those observed on-field, and ii) to evaluate the influence of varying filtering cut off frequencies on head kinematics metrics.

## MATERIALS AND METHODS

Methodological and procedural information are presented in line with the recent Consensus Head Acceleration Measurement Practices (CHAMP) recommendations ^3,18^.

### Testing Protocol

Testing was conducted via Imperial College HEAD lab’s drop rig to deliver impact to a 50^th^ percentile Hybrid III headform (also called an anthropometric testing device, ATD, in this paper) (Figure 1A). The headform was instrumented with an array of nine PCB piezoelectric accelerometers (PCB 352C23, PCB Piezotronics, Depew, New York, USA), mounted inside the headform in a 3-2-2-2 arrangement. This allowed for the measurement of both linear and rotational accelerations at the centre of mass of the headform (Figure 1B) using Padgaonkar’s method ^42^ the same as in several previous studies ^1,20,22,52,53^. The acceleration data were recorded at a 50 kHz frequency and filtered with a 4th order Butterworth low-pass filter with a 1kHz cut-off frequency. The ATD was considered the gold standard for measurement of head accelerations during impacts.

**Figure 1:**
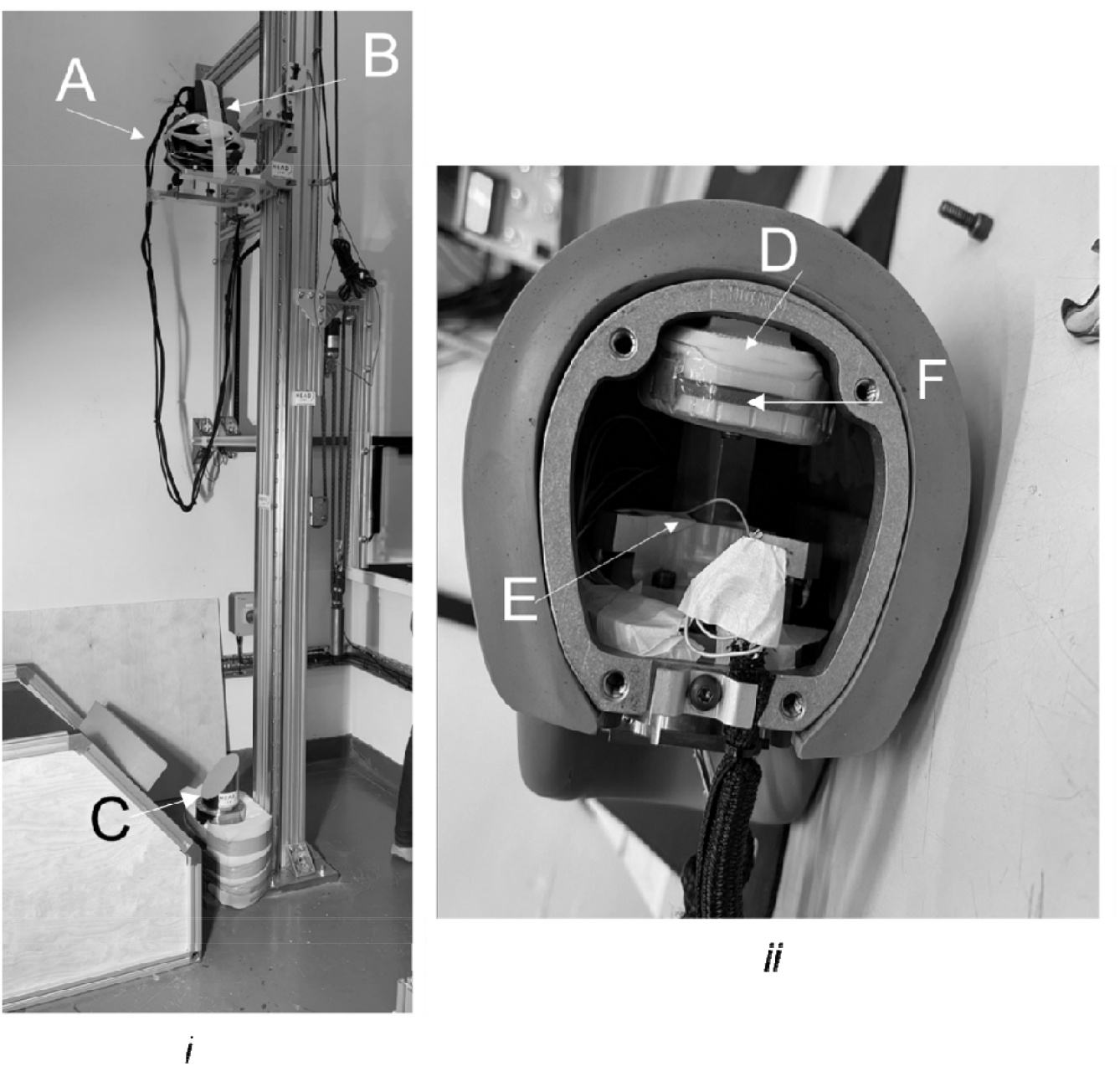
i) the testing drop rig: A = helmet, B = headform, C = Anvil. (ii) the Hybrid III headform, with mouthguard placement: D = plastic teeth, E = 9-accelerometer array, F = mouthguard.

The study compared the measurements of ATD against iMG shown in figure 1B. The mouthguard was fitted onto a plastic teeth, which was bolted into place inside and to the top of the ATD device utilising its metal casing to ensure no movement occurred. The headform was then fitted with a cycle helmet (DesignSter Lightweight Helmet) to prevent direct damage to the headform and control the acceleration magnitude and duration. The helmeted headform was lifted from the ground, and dropped onto a metal anvil angled at 45° covered with a grit 80 abrasive paper.

The headform was subjected to impacts at three locations (front, side and back) and 7 speeds (2, 3, 4, 5, 6, 7 and 8 *m/s*). These impact speeds, locations and subsequent impact durations are indicative of those observed in sports such as rugby union, boxing and other contact sports ^30,40,50^. Impacts that exceeded sensor ranges were removed from comparisons. The total number of impacts used were 82 ranging from 10 to 200 *g* in PLA. To reduce the chance of helmet failure, the helmets were only subjected to one impact to the front, side and back before being discarded, which is standard practice from previous work^1^. For consistency, checks were performed after each impact to ensure the mouthguard had not come loose before proceeding to the next impact test.

### iMG Measurement and Specifications

The iMG of the PROTECHT system contained a tri-axial accelerometer (H3LIS331DL, STMicroelectronics, Genova, Switzerland) and a tri-axial gyroscope (LSM9DS1, STMicroelectronics, Genova, Switzerland). The former was sampled at 1 *kHz* (± 400 *g*, 12-bit resolution) and the latter at 1 *kHz* (±35 *rad*.*s*^*-1*^, 12-bit resolution). For each impact, the inertial sensors collected 104 *ms* of data for the mouthguard and 2 seconds for the ATD. For both the ATD and iMG the trigger-point of the sensors was a raw linear acceleration exceeding 10 *g* in any one of the three axes. Rotational accelerations were derived from the rotational velocity time-series using a five-point stencil approximation for the mouthguard. Peak values reported were defined as the maximum numerical value of the vector-norm of the respective time-series data. A summary of the specifications and processing is outlined in Table 1.

**Table 1:**
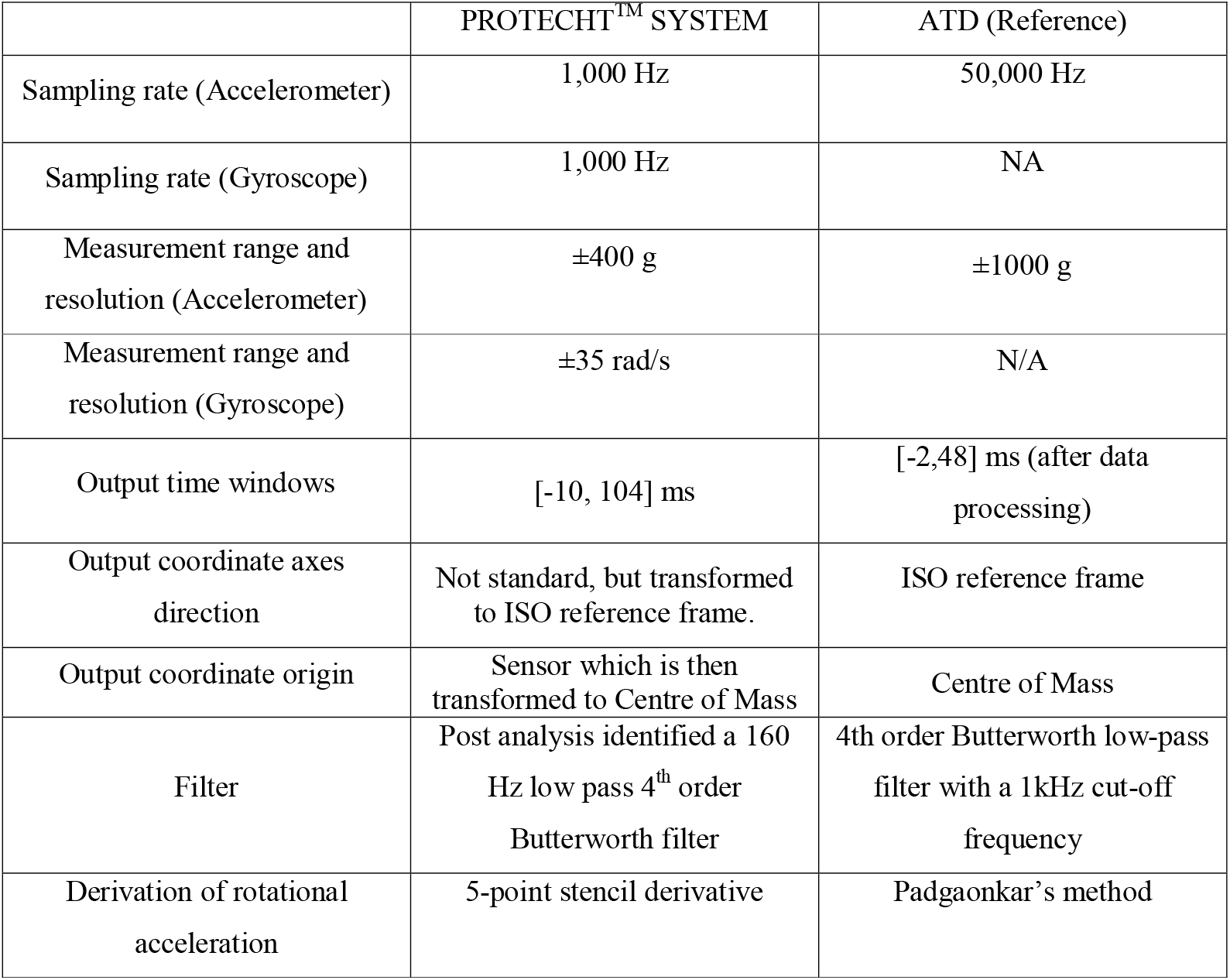
System specifications for the PROTECHT™ SYSTEM and ATD reference data.

|  | PROTECHT<sup>TM</sup> SYSTEM | ATD (Reference) |
| --- | --- | --- |
| Sampling rate (Accelerometer) | 1,000 Hz | 50,000 Hz |
| Sampling rate (Gyroscope) | 1,000 Hz | NA |
| Measurement range and resolution (Accelerometer) | $\pm 400$ g | $\pm 1000$ g |
| Measurement range and resolution (Gyroscope) | $\pm 35$ rad/s | N/A |
| Output time windows | [-10, 104] ms | [-2,48] ms (after data processing) |
| Output coordinate axes direction | Not standard, but transformed to ISO reference frame. | ISO reference frame |
| Output coordinate origin | Sensor which is then transformed to Centre of Mass | Centre of Mass |
| Filter | Post analysis identified a 160 Hz low pass 4<sup>th</sup> order Butterworth filter | 4th order Butterworth low-pass filter with a 1kHz cut-off frequency |
| Derivation of rotational acceleration | 5-point stencil derivative | Padgaonkar's method |

### Filtering Considerations

The type of filter and filter cut off frequency applied to the acceleration time-series data drastically influence head impact metric measurements ^45^. While conventions exist for the standardisation of such inputs within automotive testing – the SAE J211^46^ – no such convention exists within sports head impact measurement systems, introducing ambiguity into comparisons between systems. We approached this problem with two strategies: firstly, addressed how cut off frequency can influence peak kinematics measures, and secondly, utilised various spectral analysis measures to define the optimal cut off frequency for collected iMG data.

For the former, we compared multiple cut off frequencies to ATD collected data. The 50kHz ATD data (low pass Butterworth filtered with cut off frequency of 1000Hz) was used as a gold standard measure. The iMG from this study compared PLA, PRV and PRA outputs at various low pass, Butterworth filtered cut off frequencies of 300, 200, 100 and 50 Hz, corresponding to the most common frequencies utilised within a recent comparison of multiple iMG systems ^30^. In addition, the ATD was also down-sampled to 3200 Hz to represent the other iMGs reported within the literature ^30^, with the same cut off frequencies of 300, 200, 100 and 50Hz also applied with their corresponding output kinematic metrics compared to the 50kHz ATD data.

To establish optimal filter cut off frequency for iMG data, Fast Fourier transform (FFT) analysis was completed within Matlab R2022a (Signal Processing Toolbox; The Mathworks Inc, Natick, Massachusetts, USA) assessing the *x, y, z* and resultant components of linear acceleration, rotational velocity and rotational acceleration time-series data for both ATD and iMG systems. To provide further time-resolution to the frequency analysis of signals, continuous wavelet transformations (CWT) were also completed within Matlab (Signal Processing Toolbox; The Mathworks Inc, Natick, Massachusetts, USA), utilizing a ‘bump’ wavelet. Time-series data were zero padded to ensure the impact point lay within the cone of influence and was not subject to boundary effects.

### Finite element model of the human head

The Imperial College 3D finite element model of the human head was used to predict brain deformation during impacts. The model incorporates details of the brain anatomy from high-resolution magnetic resonance images of a healthy 34-year-old male subject. The model consists of nearly one million hexahedral elements and a quarter of a million quadrilateral elements, representing 11 tissues, including the scalp, skull, brain, meninges, subarachnoid space and ventricles. The details of the model development, mechanical properties of different tissues and the validation of model prediction of brain displacement against post-mortem human subject experiments can be found in ^21,29,53^. A non-linear transient dynamic code, LS-DYNA ^26^, was used to set up the model and solve the equations using 20 cores of a high-performance computer and 16 GB RAM.

The head model was loaded by the translational and rotational accelerations obtained from the ATD and iMG head impact data. For each element of the model, the maximum principal value of the Green-Lagrange strain tensor, which the element experienced during the impact, was determined. This quantity is called maximum principal strain (MPS) or strain hereafter. Several previous studies have shown that mechanical strain produced during head impacts can predict changes in brain tissue and vasculature ^4,12–14,16,21,38^.

### Statistical Analysis

For filter comparisons, peak resultant rotational velocity (PRV), PLA and PRA data was filtered at 300, 200, 100 and 50Hz cut off frequencies were compared using a one-way ANOVA. If main effects were present, Bonferonni adjusted post-hoc tests were administered comparing against the ATD 50kHz gold standard.

For ATD and iMG comparisons, dependant variables were PLA, PRV and PRA and MPS, as measured by the ATD and the iMG.

Scatterplots and coefficient of determination (R-squared) were calculated to assess the relationship between the ATD and iMG. R-squared values indicates the proportionate amount of variation in the response variable yY explained by the independent variables X in the linear regression model ^17^. Although a commonly used method within iMG validation methodology ^5,6^, correlation and linear regression model R squared values (interpreted in isolation) do not assess statistical agreement between measures and should not be utilised as a sole statistical test within such methodologies.

Agreement between measures was assessed using a battery of statistical tests: intraclass correlation coefficients (*ICC*), concordance correlation coefficient (*CCC*), Bland-Altman 95% limits of agreement and ordinary least products regression. Intraclass correlation coefficients measure the reliability and validity of measurements for data that have been collected as groups ^17^. For all variables, ICCs were calculated using the (3,1) convention^47^. The CCC evaluates the degree to which pairs of observations fall on the 45° line through the origin ^33^; values for linear and rotational kinematics measures and MPS were calculated. The combined *CCC* value that accounts for peak linear and rotational acceleration represented the overall iMG in-laboratory validity ^32^. The minimum validity threshold value for both *CCC* and *ICC* values is considered 0.80 ^7,32^.

Bland-Altman 95% limits of agreement analysis is a simple method to evaluate the mean difference between measurement systems, and to estimate an agreement interval within which 95% of the differences between methods falls ^2^. Bland-Altman analysis was conducted using the “blandr” package ^11^ on RStudio (RStudio, Vienna, Austria). Differences were calculated weighing towards the ATD system, meaning positive bias indicated an underestimation in the iMG. Analyses were expressed using percentage difference. Although *a priori* 95% limits of agreement are usually required for Bland-Altman analysis ^23^, there is a lack of clinically informed criteria regarding what constitutes ‘agreement’ within head impact sensors. Therefore, ordinary least products regression was also implemented, which assesses fixed and proportional bias between two measurement systems ^36^. If the 95% confidence interval for the intercept did not include 0, then fixed bias was present. If the 95% confidence interval for the slope did not include 1.0, then proportional bias was present. Ordinary least products regression analysis was completed in RStudio using the ‘lmodel2’ package ^34^.

Finally, root mean-square errors were calculated to assess the accuracy of the overall time-series data, following a modified procedure from previous research ^10^. For RMS, the peaks of corresponding impacts were first temporally aligned. They were then time-normalised based on the local minima either side of the peak value, which were identified in the ATD data. The RMS errors were also normalised (nRMS) based on the kinematics magnitude:

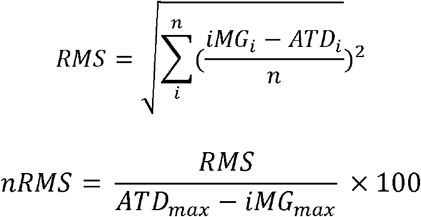

where *n* is the number of measurements, and ATDmax and ATDmin are the maximum and minimum values recorded by the ATD during the impact.

## RESULTS

### ATD vs iMG Comparisons

For ATD and iMG comparisons, 82 total impacts were completed, with a mean impact duration of 9 *ms* ranging from 6 - 18 *ms*. Due to issues with iMG pre-sampling time, only 66 trials were utilised to calculate RMSE and nRMSE. Table 2 presents descriptive statistics for PLA, PRV, PRA and MPS data for all 82 impacts, which confirms a large range of kinematics achieved in the tests.

**Table 2:** Descriptive statistics of all impacts for ATD and iMG.

|  | PLA (g) |  | PRV (rad/s) |  | PRA (rad/s/s) |  | MPS |  |
| --- | --- | --- | --- | --- | --- | --- | --- | --- |
|  | ATD | iMG | ATD | iMG | ATD | iMG | ATD | iMG |
| *n* | 82 | 82 | 82 | 82 | 82 | 82 | 81 | 81 |
| Median | 103.0 | 106.2 | 30.1 | 28.6 | 6069 | 5839 | 0.189 | 0.187 |
| Mean | 99.2 | 102.2 | 29.1 | 28.2 | 6116 | 5627 | 0.187 | 0.184 |
| St Dev | 40.4 | 43.8 | 9.48 | 8.56 | 2366 | 2011 | 0.06 | 0.06 |
| Minimum | 16.0 | 14.5 | 11.3 | 11.9 | 1331 | 1381 | 0.058 | 0.059 |
| Maximum | 175.4 | 191.4 | 52.0 | 42.8 | 11021 | 8851 | 0.280 | 0.287 |

Representative examples of the time-normalised vector norms of the measured time-series data are presented in Figure 2. Solid and dashed curves represent ATD and iMG time-series data (respectively) for impacts at 2, 4 and 6 m/s for linear acceleration, rotational velocity and rotational acceleration.

**Figure 2:**
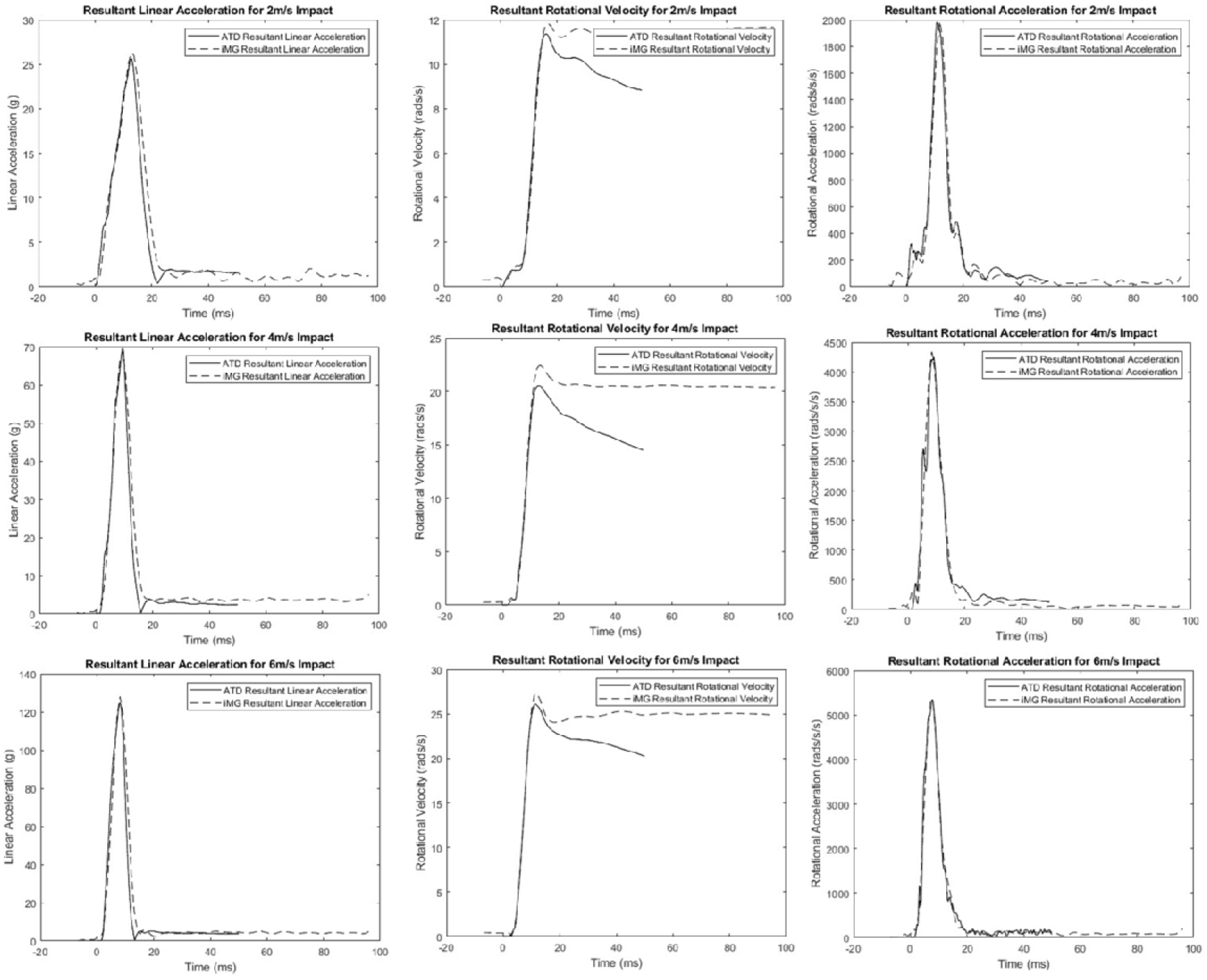
Examples of the ATD (solid line) and iMG (dashed line) measured head kinematics. The curves are synched using the peak values and then used to calculate RMS and NRMS errors. Column 1 represents linear acceleration curves, column 2 rotational velocity curves, and column 3 rotational acceleration curves. Row 1 represents a 2m/s impact, row 2 a 4 m/s impact, and row 3 a 6 m/s impact.

Figure 3 presents scatter plots and R^2^ values for PLA (R^2^ = 0.9), PRV (R^2^ = 0.93), PRA (R^2^ = 0.95) and MPS (R^2^ = 0.94) for the iMG system compared to the ATD reference system. Statistical results are presented in table 3, which report the ICC values, CCC values, RMSE, nRMSE and Bland-Altman statistics for PLA, PRV and PRA for the iMG compared to the ATD reference system. As a short summary, all ICC values indicated excellent reliability; all CCC values indicated substantial agreement and exceeded the minimum required value of 0.8^32^ and minimal bias was detected within Bland-Altman analysis. Bland Altman plots, for both absolute and % difference measures, are presented in figure 4.

**Table 3:** Agreement statistics for biomechanical metrics: Intraclass correlation coefficients (ICC), Concordance correlation coefficients (CCC), RMSE, nRMSE and Bland Altman Statistics for PLA, PRV and PRA in the ATD and iMG for the subset of 66 impacts.

| | CCC<br>(95% CI) | ICC<br>(95% CI) | Mean<br>Relative<br>Error % | RMSE<br>(± SD) | nRMSE<br>(±SD; %) | Bland Altman (% Difference) | | |
| --- | --- | --- | --- | --- | --- | --- | --- | --- |
|  |  |  |  |  |  | Bias (95%<br>CI) | Lower<br>Limit | Upper<br>Limit |
| **PLA (g)** | 0.989<br>0.984-0.992 | 0.992<br>0.987-0.995 | 2.20% | 6.73<br>(±3.23) | 6.45<br>(±1.96) | -2.16%<br>(-4.51% -<br>0.19%) | -14.43% | 10.10% |
| **PRV (rad/s)** | 0.970<br>0.953-0.980 | 0.971<br>0.955-0.981 | -0.70% | 2.18<br>(±1.13) | 7.57<br>(±2.71) | 0.69%<br>(-2.07% -<br>3.46%) | -13.74% | 15.12% |
| **PRA**<br>**(rad/s<sup>2</sup>)** | 0.945<br>0.921-0.962 | 0.965<br>0.946-0.977 | -6.20% | 375<br>(±209) | 6.13<br>(±2.37) | 6.16%<br>(3.31% - 9.02%) | -8.72% | 21.04% |
| **MPS** | 0.970<br>0.955-0.981 | 0.972<br>0.957-0.982 | 1.30% | N/A | N/A | 1.35%<br>(-1.59% -<br>4.29%) | -13.85% | 16.59% |

**Figure 3:**
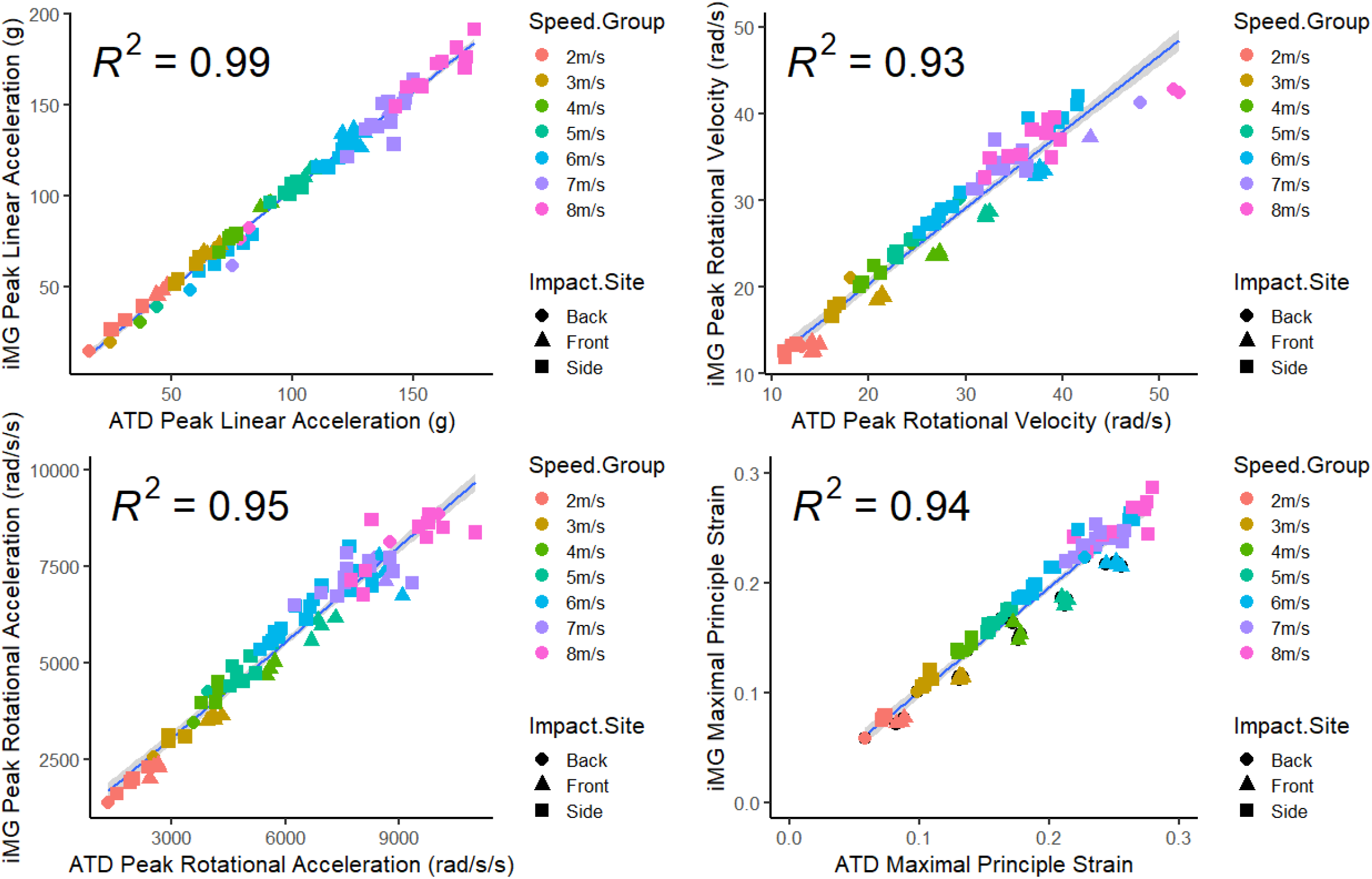
Scatter plots for peak linear acceleration, peak rotational velocity, peak rotational acceleration and maximal principal strain, measured by the ATD and iMG. Linear regression trendline (±95% CI on line) and R squared values displayed on each graph.

**Figure 4.**
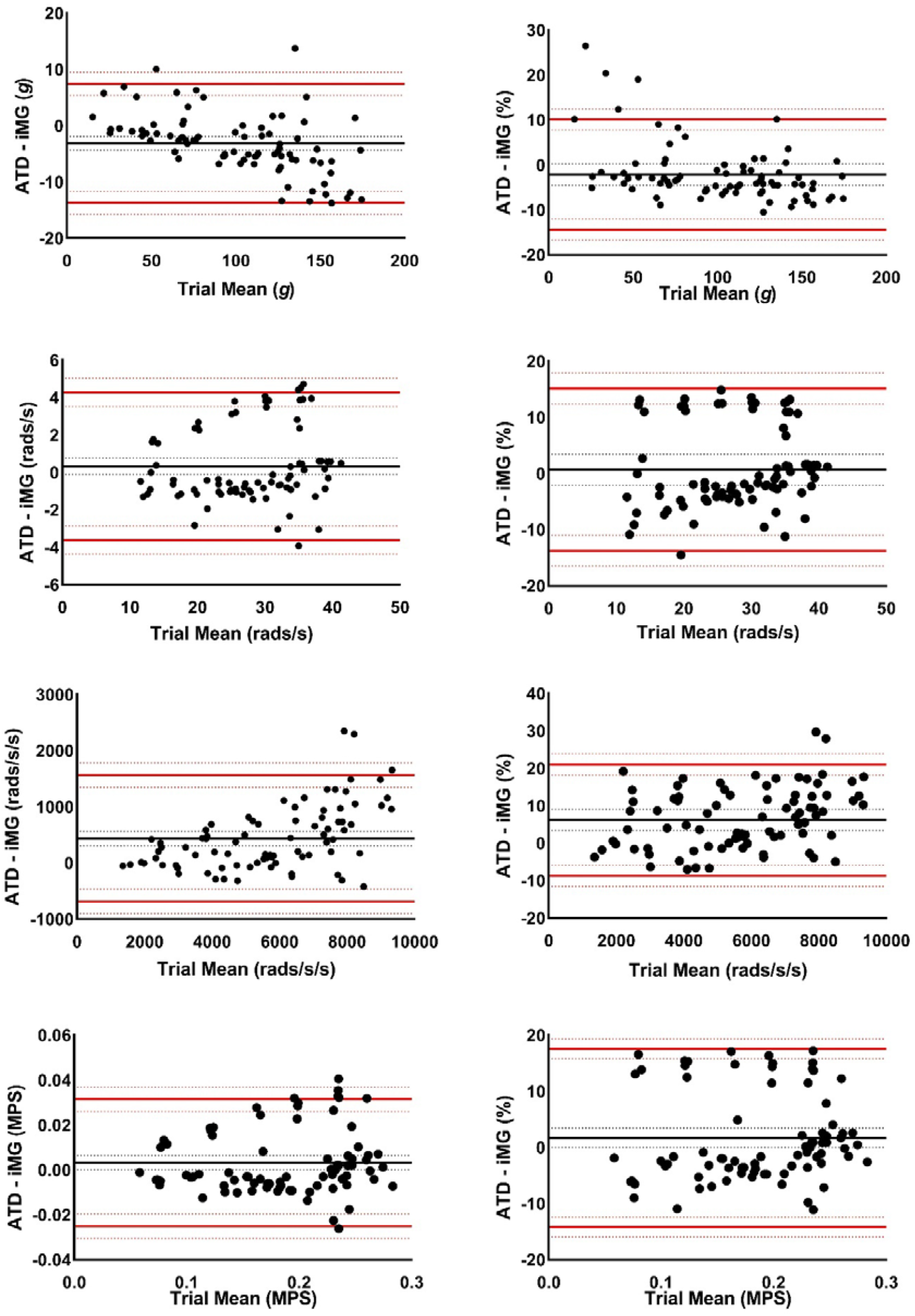
Bland Altman Plots for 95% Limits of agreement between ATD (left column) and iMG (right column) for PLA (1^st^ row), PRV (2^nd^ row), PRA (3^rd^ row) and MPS (4^th^ row).

Ordinary least products regression analysis found evidence of fixed (intercept = 3.707; 95% CI 1.37 – 5.99) and proportional (slope = 0.93; 95%CI 0.91 – 0.95) bias for PLA; and evidence of fixed (intercept = 1.15; 95% CI 1.09 – 1.21) and proportional (slope = -400; 95% CI -726 – -89) bias within PRA. There was no evidence of fixed (intercept = -0.5; 95% CI 0.98 – 1.09) or proportional (slope = 1.03; 95% CI 0.98 – 1.08) bias for PRV. There was no evidence of fixed (intercept = -0.0000683; 95% CI -0.0108 – 0.00892) or proportional (slope = 1.021; 95% CI: 0.9688 – 1.0762) bias for MPS.

### Effects of the Filter Cut Off Frequency – Downsampled ATD Data

For filter cut off frequency comparisons, 82 impacts were utilised. For ATD data that was down sampled to 3200Hz, there was a significant main effect of filter condition for PLA (F_(4,445)_ = 22.158, p < 0.001, η^2^ = 0.166) and PRA (F_(4,445)_ = 45.455, p < 0.001, η^2^ = 0.290) but not PRV (F_(4,445)_ = 0.570, p =0.684, η^2^ = 0.005). Post hoc comparisons and descriptive statistics are shown in Figure 5.

**Figure 5.**
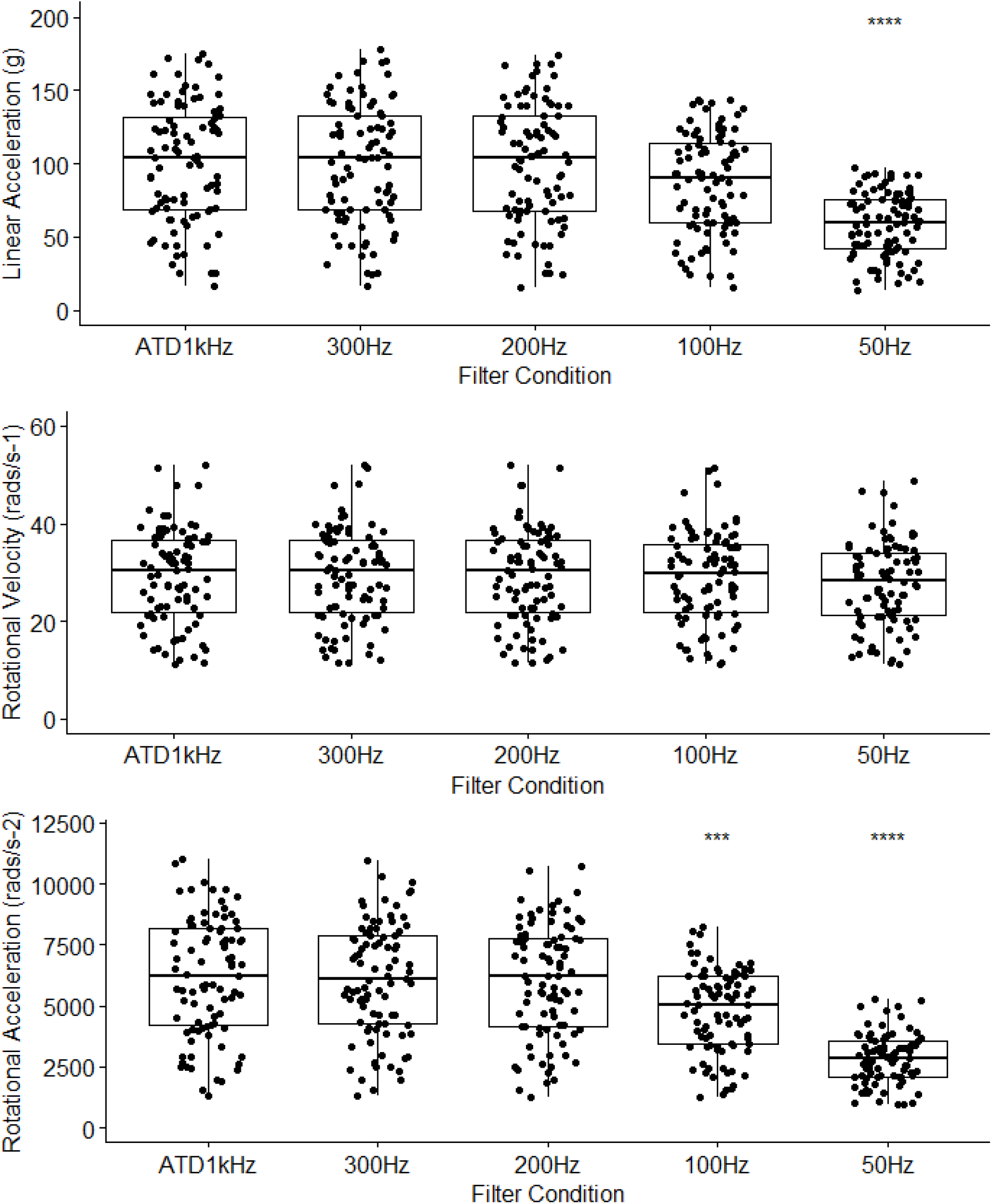
Box plots comparing filter conditions for PLA, PRV and PRA for downsampled ATD data. * = p<0.05, ***==p<0.001, **** = p<0.0001; all comparisons made against reference category of ATD 1kHz.

### Effects of the Filter Cut Off Frequency – iMG Data

For iMG data, there was a significant main effects for filter condition for PLA (F_(5,378)_ = 14.203, p < 0.001, η^2^ = 0.158) with only the 50Hz condition significantly underestimating the reference ATD value by 40.7 *g* (95%CI = 19.2 to 62.2; p<0.001). There was a significant main effect for filtered condition on PRA (F_(5,378)_ = 34.398, p < 0.001, η^2^ = 0.313), with the 50Hz condition significantly underestimating the reference ATD value by 4470 rads/s^2^ (95% CI = 3395 to 5545; p < 0.001). There was no significant main effect for filter condition for PRV (F_(5,378)_ = 0.283, p = 0.923, η^2^ = 0.004). Post hoc comparisons and descriptive statistics are shown in Figure 5 and Table 4 by breakdown of linear magnitude intensity buckets.

**Table 4.** Descriptive stats for mean PLA, PRV and PRA for iMG data unfiltered, and filtered at 50-300 Hz cut off frequencies compared to the gold standard ATD data set using a 1000 Hz cut off frequency.

| Impact Magnitude | Filter CutOff Frequency | Mean PLA (g) | % Difference from ATD | Mean PRV (rads/s) | % Difference from ATD | Mean PRA (rads/s/s) | % Difference from ATD |
| --- | --- | --- | --- | --- | --- | --- | --- |
| 0-50g | ATD | 37.8 | NA | 13.065 | NA | 2250 | NA |
|  | Raw iMG | 36 | 4.82% | 12.9 | 1.16% | 2373 | 5.47% |
|  | 300Hz iMG | 35.4 | 6.36% | 12.9 | 1.32% | 2090 | 7.11% |
|  | 200Hz iMG | 35.2 | 6.85% | 12.9 | 1.30% | 2119 | 5.82% |
|  | 100Hz iMG | 32 | 15.35% | 13 | 0.71% | 1795 | 20.22% |
|  | 50Hz iMG | 22.9 | 39.43% | 13 | 0.51% | 1251 | 44.40% |
| 50-100g | ATD | 76 | NA | 27 | NA | 5066 | NA |
|  | Raw iMG | 84.5 | 11.29% | 26.2 | 3.12% | 5869 | 15.85% |
|  | 300Hz iMG | 83.2 | 9.48% | 26.1 | 3.41% | 5230 | 3.24% |
|  | 200Hz iMG | 82.6 | 8.79% | 26.1 | 3.57% | 4969 | 1.91% |
|  | 100Hz iMG | 73.9 | 2.68% | 26.1 | 3.54% | 4101 | 19.05% |
|  | 50Hz iMG | 48.6 | 36.00% | 25.5 | 5.81% | 2399 | 52.65% |
| 100g-150g | ATD | 127 | NA | 31.5 | NA | 7182 | NA |
|  | Raw iMG | 139.9 | 10.18% | 31.2 | 1.22% | 8016 | 11.61% |
|  | 300Hz iMG | 137.5 | 8.28% | 30.9 | 1.98% | 7081 | 1.41% |
|  | 200Hz iMG | 136 | 7.13% | 31 | 1.68% | 6771 | 5.72% |
|  | 100Hz iMG | 117.6 | 7.40% | 30.8 | 2.20% | 5263 | 26.72% |
|  | 50Hz iMG | 74.9 | 41.00% | 29.6 | 6.07% | 2884 | 59.84% |
| 150g+ | ATD | 167.8 | NA | 37.9 | NA | 9728 | NA |
|  | Raw iMG | 204.4 | 21.77% | 37.5 | 0.99% | 11876 | 22.08% |
|  | 300Hz iMG | 198.7 | 18.42% | 37.4 | 1.42% | 10544 | 8.39% |
|  | 200Hz iMG | 192.8 | 14.88% | 37 | 2.38% | 9150 | 5.94% |
|  | 100Hz iMG | 162.2 | 3.36% | 36.5 | 3.72% | 6289 | 35.35% |
|  | 50Hz iMG | 101.1 | 39.76% | 35.4 | 6.57% | 3417 | 64.87% |
| Total | ATD | 103.827 | NA | 28.787 | NA | 6198 | NA |
|  | Raw iMG | 116.302 | 12.02% | 28.26 | 1.83% | 7077 | 14.18% |
|  | 300Hz iMG | 114.134 | 9.93% | 28.106 | 2.37% | 6273 | 1.21% |
|  | 200Hz iMG | 112.707 | 8.55% | 28.108 | 2.36% | 5910 | 4.65% |
|  | 100Hz iMG | 98.042 | 5.57% | 27.988 | 2.78% | 4617 | 25.51% |
|  | 50Hz iMG | 63.04 | 39.28% | 27.115 | 5.81% | 2607 | 57.94% |

### Optimal Filter Cut Off Frequency

FFT and CWT outputs determined the use of a 160 *Hz* Low-pass 4^th^ order Butterworth filter for iMG data. This universal cut off frequency best represents previous work^31^ and the interpretation of the FFT outputs. Example FFT and CWT graphs for 2 m/s, 4 m/s and 6 m/s impacts, and the MATLAB code used to generate them, can be seen in supplementary materials. Optimal cut off frequency was chosen as the point at which the FFT graph appears to visually plateau following the large spike in the early frequencies. The representative graphs show this point with a red line. This point varied across impacts with different speeds, with linear acceleration values varying between 130Hz and up to 200Hz for the higher speed impacts. 160Hz was chosen as an optimum cut off frequency value that best represented all measured impacts. It is acknowledged that the point of inflection/visual plateau within the rotational velocity curves falls earlier than 160Hz for the majority of impacts, but as a higher cut off frequency did not appear to introduce additional noise into the velocity signal, 160Hz was also used. Further investigation identified that the impact velocity affected the frequency domain to individual axis components seen in the CWT graphs, and though a universal filter elicited close agreement with the ATD, a future refined approach of using a moving cut-off frequency dependant on the impact magnitude and change to the frequency domain should be explored.

## DISCUSSION

One aim of this study was to compare the ATD reference system against an iMG system under large range of impact conditions that represented those observed on field, taking current comparison studies one step further. The results showed good agreements and strong positive correlations between the head kinematics measured by the iMG and ATD. In addition, predicted brain strains based on ATD and iMG systems showed good agreement. As the second aim, multiple low-pass filter cut off frequencies were compared to assess their influence on peak kinematics metrics. This analysis showed that low cut off frequencies of 100 and 50Hz significantly lowers peak linear and rotational accelerations, but not rotational velocity. A cut off frequency of 160 Hz was determined suitable for the large range of head kinematics achieved in our experiments.

The measured iMG acceleration time-series data had a total *CCC* value of 0.970 when compared to the ATD reference measurement system, under conditions that expose the iMG device to PLA ranges of 10-200 g and impact durations of 6 - 18 *ms*. This is the first time from which an iMG device has been tested under these conditions, which produce PLAs almost double those reported in previous literature. The reported *CCC* value have met the threshold (>80%) outlined by Kieffer et al.^32^ in their two-phase approach to quantifying head impact sensor accuracy, indicating the iMG would be suitable to complete the second stage of evaluation measuring on-field kinematics. The total *CCC* value reported in the current study is similar to those reported by previous literature evaluating a cohort of instrumented mouthguards ^30^. The authors reported total *CCC* values of 0.953-0.983 from the highest performing iMG systems when compared with the ATD reference system. A previous validation reported CCC values of 0.967 of an older version of the PROTECHT system iMG

^31^. However, testing conducted by both aforementioned studies utilised an alternative experimental set up: the testing conducted by Jones et al.^30^ used a pendulum impactor on a bareheaded ATD headform, and the testing reported by Jones & Brown^31^ used a pneumatic linear impactor on a helmeted ATD headform, with both studies achieving PLAs up to 100 *g* due to equipment limitations. PLAs of over 100 *g* have been reported in sports such as boxing, American football and rugby ^9,25,30,39^. Such high acceleration magnitudes have an increased chance of causing traumatic brain injuries, although it should be noted that increased strain magnitudes of brain tissues are predominantly due to the increased rotational kinematics accompanying high linear kinematics^44^. As the reconstruction and modelling of concussive impacts could enable researchers to better investigate causal mechanisms of injury, the validity and reliability of on field kinematics inputs is vital, requiring iMGs to be validated within a large range of kinematics likely to occur on the field as done in this study.

The measured iMG and ATD data were positively correlated, with R-squared values of 0.99 for PLA, PRV and PRA and relative errors of 2.2%, -0.7% and -6.2% for PLA, PRV and PRA. Intraclass correlation coefficient (ICC) values of 0.992, 0.971 and 0.965 for PLA, PRV and PRA respectively met the minimum acceptability for reliability and validity measures of >0.80 ^7^. The findings of this study are consistent with previous research that has compared the accuracy of other instrumented mouthguards for collision sports, with Greybe et al., ^24^ reporting *ICC* values of 0.95 and 0.99 for PLA and PRV respectively and those reported by Jones and Brown.^31^ with *ICC* values of 0.96 and 0.99 and 0.95 for PLA, PRV and PRA respectively.

Ordinary least products regression analysis showed fixed and proportional bias for both linear and rotational acceleration. To some degree this is evident with the Bland-Altman plots reported in figure 4, where there is slope evidence within both the linear and angular acceleration plots for absolute values. Proportional bias could suggest that as the magnitude of impact increase, the amount of variability within the agreement of the two methods also increases. However, when analysed via percentage difference, data conformed to a homoscedastic distribution, suggesting the magnitude of error only increases in line with the magnitude of impact. Such approaches were also used by previous mouthguard evaluations^30^. Bland-Altman analysis showed slight overestimation of PLA within the iMG of 2.2% and larger underestimation of PRA by 6.16%. Limits of agreement ranged between -14.4 to 10% for PLA, between -13.7 to 15% for PRV and between -8.7 to 21% for PRA. There are currently no clinically meaningful criteria for what represents acceptable agreement with iMGs. However, these results were comparable to previous mouthguard validations, where the top performing mouthguard reported 31.7% and 29.7% limits agreement for PLA and PRA respectively ^24,30^.

It is also important to evaluate the accuracy of the overall shape of the time-series data as many in-house machine learning classification processes utilise time-series feature analysis^10^. To that end, RMS and NRMS errors of the time-series data were calculated utilising a modified methodology of Camarillo et al. ^10^. In that study, the RMS error was calculated over a 25-ms period, centred on the impact maximum, which was assumed to capture the relevant portion of the impact trace. In the present study, we calculated the RMS (and NRMS) error over the impact portion of the measured trace. The mean RMS (and NRMS) error for the linear acceleration and rotational velocity was 6.7 ± 3.2 g (6.5 ± 1.9%) and 2.2 ± 1.1 rad/s (2.6 ± 2.7%), respectively. These errors are comparable with – and on a normalised basis, lower than - those reported by Camarillo et al^10^, where the mean RMS (and NRMS) errors for the linear acceleration and rotational velocity were 3.9 ± 2.1 g (9.9 ± 4.4%) and 1.0 ± 0.8 rad/s (10.4 ± 9.9%), respectively. The small RMS error between the measured timeseries data (both linear acceleration and rotational velocity) indicates high comparability of the overall shape of the waveforms measured by the two sensor systems.

Maximal principal strain measures predicted by inputs from ATD or iMG showed high levels of agreement, with a CCC value of 0.971, an ICC of 0.972 and mean relative error of 1.3%. Bland-Altman 95% limits of agreement analysis reports a bias of 1.35% with limits ranging between -13.85% and 16.59%, and ordinary least products regression reported no fixed or proportional bias. As previously discussed, although Bland-Altman analysis usually requires an *a priori* set level of agreement between the two measurements, there is currently no set level of agreement informed by clinical measures within the brain modelling literature. The greatest relative error for MPS occurred in the test with the largest difference in rotational acceleration between the ATD and iMG systems. The linear acceleration had no effect on the MPS, in agreement with the findings of previous research^15^. Hence, an accurate measurement of rotational kinematics is important for an accurate prediction of the MPS value. Only one previous study has compared the predicted brain strain across ATD and iMG input kinematics, where mean relative errors ranged between 7.5% and 8.9% for various iMGs^35^, compared to the 1.3% relative error in the current study.

Filter comparisons show that the choice of cut off frequency can have a profound effect on impact kinematics. Chosen to reflect filtering techniques from iMGs used in previous research^30^, the choice of cut off frequency significantly influenced linear and rotational acceleration, both with large effect sizes (η^2^ = 0.166 and η^2^ = 0.290, respectively).

Specifically, a 50 Hz cut off frequency significantly reduced PLA and PRA when compared to the ATD 1kHz gold standard, and a 100 Hz cut off frequency significantly reduced rotational accelerations compared to the gold standard measure. Though not significant for PLA, there were differences reported when using 100 Hz cut off frequency at a number of impact magnitude ranges as seen in Figure 2. Filter cut off frequencies of 300Hz and 200Hz did not significantly attenuate impact kinematics. Given the importance of rotational kinematics for brain strain predictions, the use of iMG data with low cut off filters could lead to an underestimation of strain predictions. While it is acknowledged that the choice of cut off frequency is likely to be related to the characteristics of impact, a lack of consensus on impact data treatment could lead to ambiguity when impact kinematics from different systems are compared.

A limitation of the current study is the placement of the iMG at the top of ATD, as the headform used could not facilitate for placement within the ‘jaw’, similar to procedures of previous studies ^24^. While a jaw placement would emulate *in vivo* conditions to a greater extent, it has been noted that maintaining the position of the ‘jaw’ within a headform is problematic due to no active mandible contraction ^35^. Regardless, the iMG accelerations were still transformed to the ATD headform centre of mass to match the reference sensors. Furthermore, rigid coupling of the iMG to the ATD also allowed for the repeatable testing of impacts particularly those that produced large accelerations. Future research shall address on field testing validity and reliability, and player and practitioner feasibility and useability to allow for comparisons with other iMG systems^30^.

This is the first study to measure the agreement between the ATD reference system and an iMG system for impacts producing PLAs over 100 *g* and impact durations that are compatible with those observed on field in sport with reported *CCC* values of 0.970. The ability to validate under conditions that reflect on field kinematics in terms of measured impact magnitudes and durations is essential to ensure the values reported on field are correct. In this study the iMG system was comparable to that of a gold standard measure, measuring impacts magnitudes up to 200 *g* and impact durations ranging from 6 -18 *ms*. As such, the iMG system can be utilised by applied practitioners to validly and reliably measure head impact kinematics within real world game scenarios. The methods presented here can be used in future studies to validate iMG systems under laboratory conditions that reflect those observed on field to provide confidence in results and to determine optimal cut off frequencies for reliable data processing and reporting.

## Data Availability

All data produced in the present work are contained in the manuscript

## Funding

Independent laboratory testing and brain simulations completed at Imperial College London HEAD lab were funded by Sport & Wellbeing Analytics (SWA).

## Conflict of Interests

CJ, KA and KN are all employed by Sports & Wellbeing Analytics (SWA), manufacturers of the PROTECHT mouthguard. ML is part of the SWA executive board. All data testing and brain simulations were completed independently by MG, XY and CB at Imperial College London and the subsequent analyses were completed by CJ, KA and KN. Statistical analyses and integrity of results were confirmed by an independent researcher (SA) who is not employed by, or receives no benefit from, SWA. The results of the current study do not constitute endorsement of the product by the authors or the journal.

## Data Availability Statement

Data is available upon request by emailing the lead author, CJ.

## Ethical approval

Not applicable

