## Supplemental Figures for "MEASUREMENT EFFICIENCY OF AN INSTRUMENTED MOUTHGUARD UNDER A LARGE RANGE OF HEAD ACCELERATIONS AND THE EFFECTS OF FILTERING"

**MEASUREMENT EFFICIENCY OF AN INSTRUMENTED MOUTHGUARD AND THE EFFECTS OF CUTT OFF FREQUENCY**

### Supplementary Material 1

Example FFT plots utilised to establish optimal cut off frequency. The red line indicates the approximate transition from high to low magnitude components (plateau).

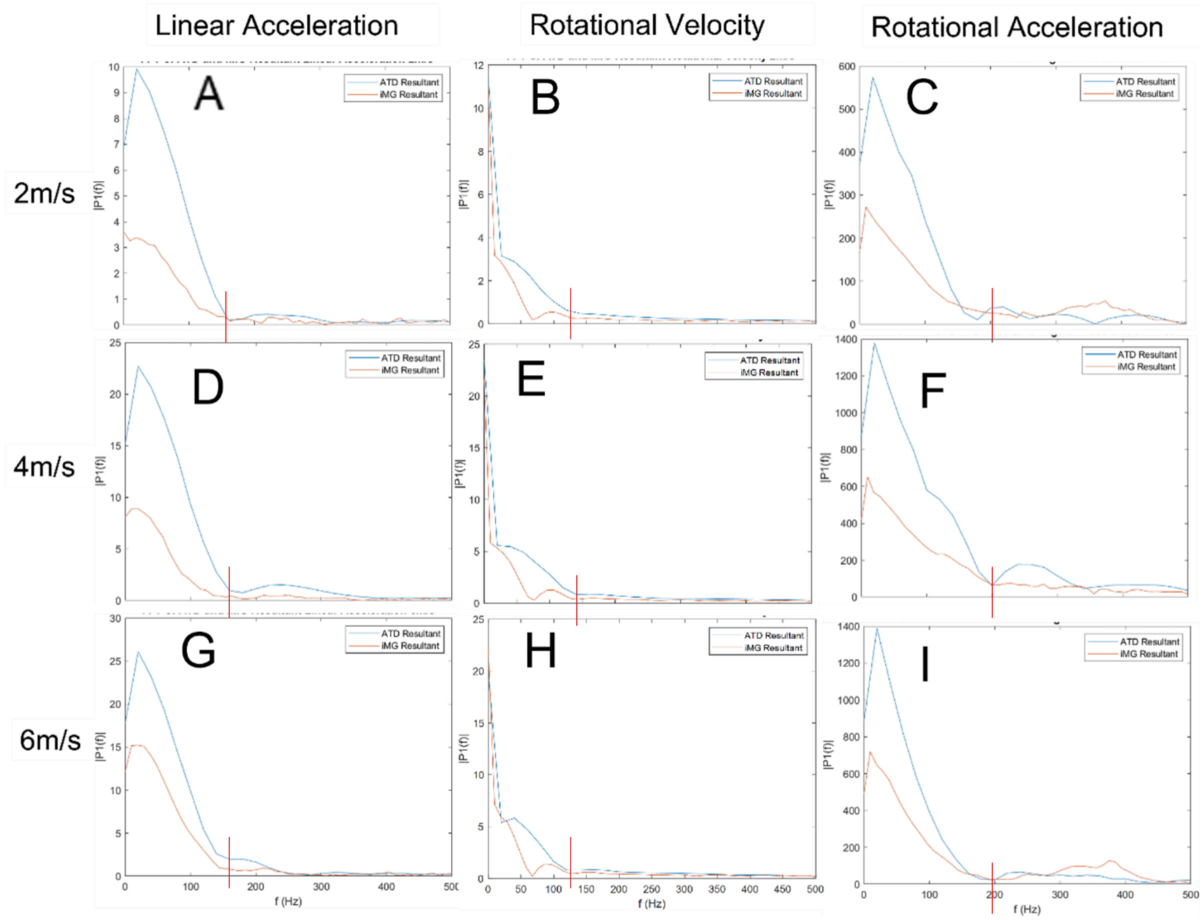

Christopher M. Jones[1*], Kieran Austin[1], Simon Augustus[2], Kieran Jai Nicholas[1], Xiancheng Yu[3], Claire Baker[3], Mike Loosemore[4,5], Mazdak Ghajari[3]

**Supplementary Material 2**

MATLAB script used to produce FFT graphs.

```
%READ DATA%
%x tests split into x corresponding excel sheets for both mouthguard and headform.

no_tests = 67

for n = 1:no_tests
    mgData(:,:,n) = xlsread('mgWaveforms.xlsx',n);
    %hfData(:,:,n) = xlsread('hfWaveforms.xlsx',n);
end
%%
% clearvars allows to run through script again without having to reimport
% all data

clearvars -except hfData mgData

%Extracts linear acceleration data for MG for every test%
%MG data converted to m/s to allow for translation to C.O.G.
for m = 1:no_tests
    mgT(:,m) = mgData(:,19,m);
    mgLinX(:,m) = (mgData(:,5,m));
    mgLinY(:,m) = (mgData(:,6,m));
    mgLinZ(:,m) = (mgData(:,4,m));
end

%% Data for Linear Acceleration FFT (replicated for Rotational
Velocity/Acceleration

%Specify individual trial from overall dataset
LinX = mgLinX(:,[1 2 3]);
LinY = mgLinY(:,[1 2 3]);
LinZ = mgLinZ(:,[1 2 3]);
AllLin = [LinX LinY LinZ]; %Combine into one dataset

%FFT Inputs
Fs = 1000;              % Sampling frequency
T = 1/Fs;               % Sampling period
L = length(LinX);               % Length of signal
t = (0:L-1)*T;          % Time vector

for k = 1: width(AllLin)
    y1(:,k) = fft(AllLin(:,k));
    P2 = abs(y1(:,k)/L);
    P1(:,k) = P2(1:L/2+1);
    P1(2:end-1,k) = 2*P1(2:end-1,k);
    f(k,:) = Fs*(0:(L/2))/L;

 end
```

Christopher M. Jones[1*], Kieran Austin[1], Simon Augustus[2], Kieran Jai Nicholas[1], Xiancheng Yu[3], Claire Baker[3], Mike Loosemore[4,5], Mazdak Ghajari[3]

**Supplementary Material 3**

Example CWT plots utilised to establish optimal cut off frequency. A, B and C represent *x*, *y* and *z* components of an impact at 2 m/s respectively; D, E and F represent *x*, *y* and *z* components of an impact at 4 m/s respectively; and G, H and I represent *x*, *y* and *z* components of an impact at 6 m/s respectively.

Linear acceleration plots

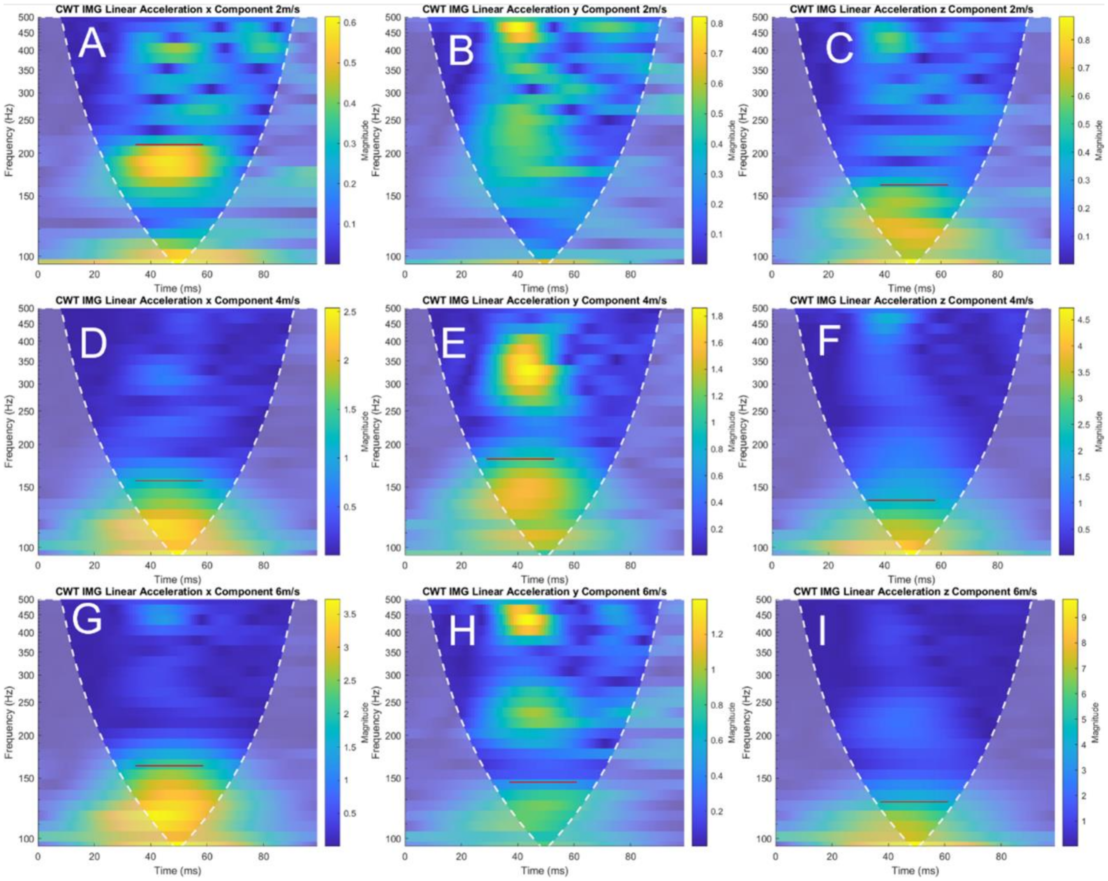

Christopher M. Jones[1*], Kieran Austin[1], Simon Augustus[2], Kieran Jai Nicholas[1], Xiancheng Yu[3], Claire Baker[3], Mike Loosemore[4,5], Mazdak Ghajari[3]

### Rotational velocity plots

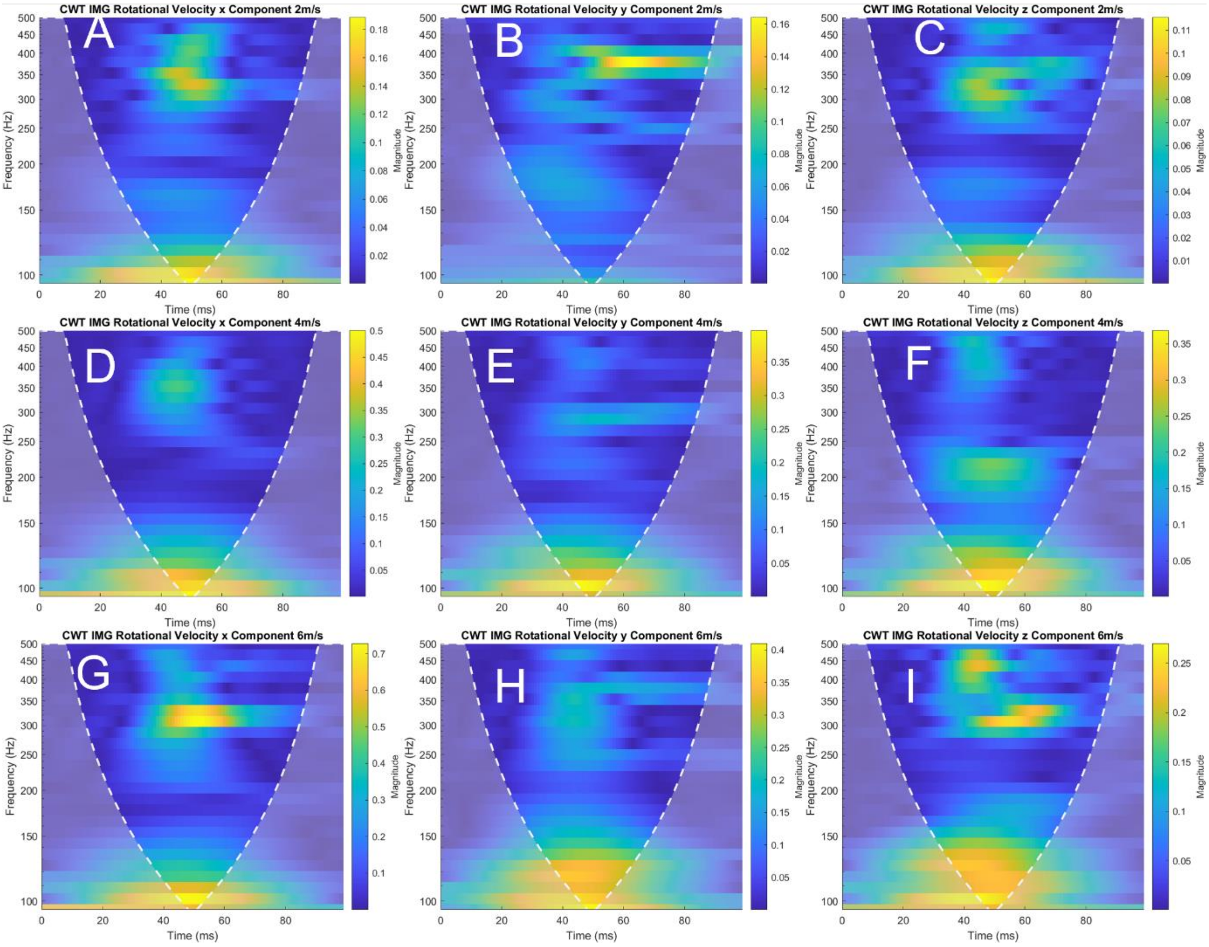

Rotational acceleration plots

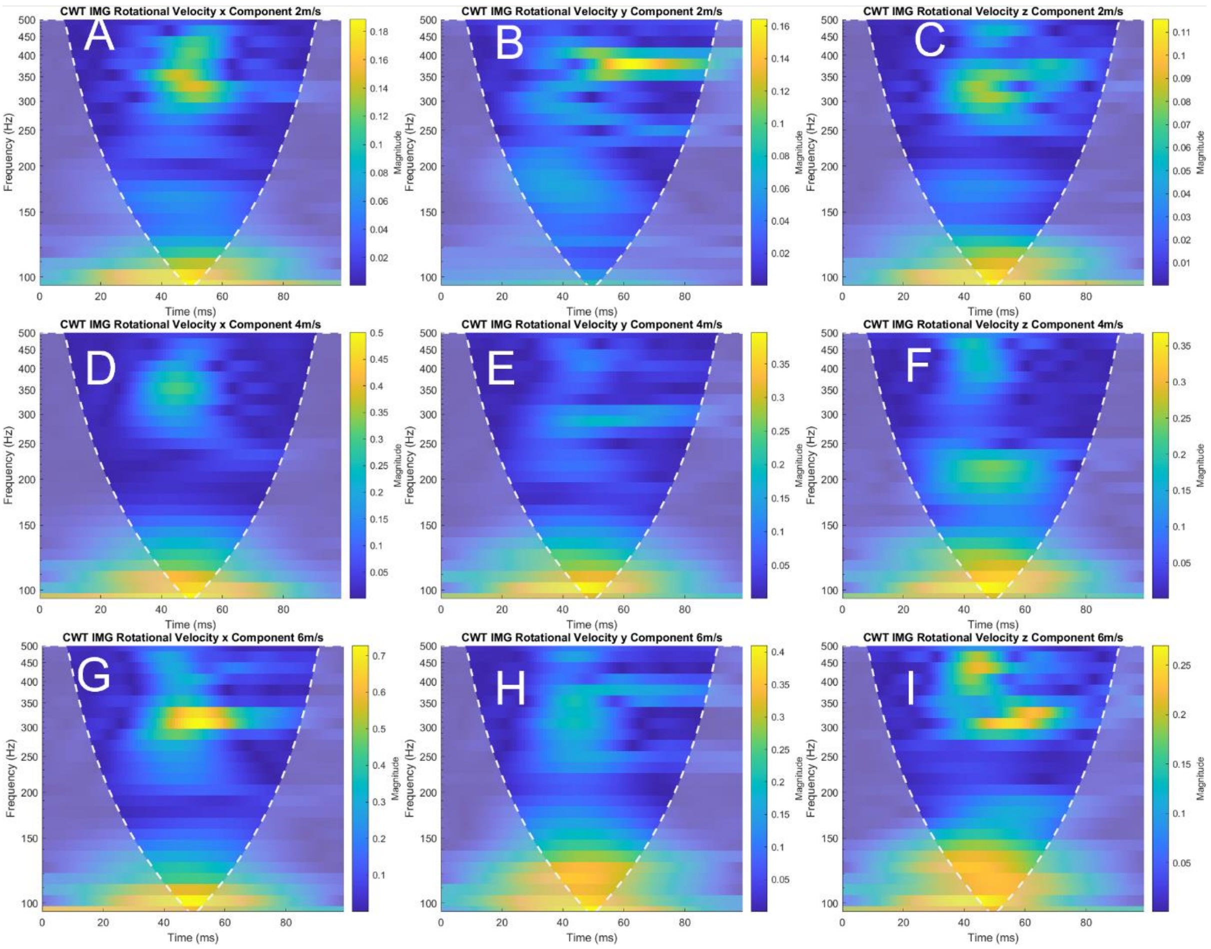


CWT Interpretation

Continuous Wavelet Scalograms provide a representation of the magnitude of a time-series signal in frequency and time domains. This enables a time localisation of high magnitude frequency components that cannot be seen in FFT visual representations. In the above figure, the dotted white lines represent the cone of influence – the area outside of this represents the region where the wavelet power spectra are distorted due to the influence of the end point of finite time signals. Time-series signals were zero padded to ensure the impacts occur within the cone of influence.

The colours represented on the graph show the magnitudes of frequency components of the signal, with the lighter (more yellow) colours showing higher magnitudes. All 'bands' of yellow appeared to be occurring around the time of impact, indicating that the higher magnitude components were most likely true signals. In some cases, for instance in graphs B, E and H, there are higher magnitude components occurring in the higher frequencies, such as above 400Hz. These high frequency components are likely to be noise, due to the area of low magnitude components between them and the lower frequency components produced by impact.

The red lines on the CWT plots indicate where the cut off frequency should be applied. This was decided as the frequency at which the magnitude of the signal components begins to return to the 'baseline' value of the surrounding signal components.

Christopher M. Jones[1*], Kieran Austin[1], Simon Augustus[2], Kieran Jai Nicholas[1], Xiancheng Yu[3], Claire Baker[3], Mike Loosemore[4,5], Mazdak Ghajari[3]

**Supplementary Material 4**

MATLAB script used to produce CWT plots.

```
%CWT Loop
% Splits Master Excel document with multiple (no_tests) sheets into single
% array

no_tests = 64

for n = 1:no_tests
    mgData(:,:,n) = xlsread('Master_VT.xlsx',n);
end

%Extracts linear acceleration data for MG for every test%
for m = 1:no_tests
    %mgT(:,m) = mgData(:,19,m); Time
    mgLinX(:,m) = (mgData(:,1,m))*9.81;
    mgLinY(:,m) = (mgData(:,2,m))*9.81;
    mgLinZ(:,m) = (mgData(:,3,m))*9.81;
    mgLinRes(:,m) = sqrt((mgLinX(:,m)).^2 + (mgLinY(:,m)).^2 + (mgLinZ(:,m)).^2);
end

%% Zero Pad Data - To ensure impact point is within cone of influence, final 30ms
of trial is removed, and 30ms of 0 value data is added to start of trial.

% Padding for IMG Data
mgLinX = mgLinX(1:70,:);
mgLinY = mgLinY(1:70,:);
mgLinZ = mgLinZ(1:70,:);
w = width(mgLinX)
zero=zeros(30,w);
mgLinX = cat(1,zero,mgLinX);
mgLinY = cat(1,zero,mgLinY);
mgLinZ = cat(1,zero,mgLinZ);


%% CWT Loop Linear X

Fs = 1000 % Specify sampling frequency

for m = 1:no_tests
    figure;
    cwt(mgLinX(:,m),'bump',Fs, FrequencyLimits = [0 500]);
    title(sprintf('CWT Linear X Trial%d'),m);
    filename = m;
    saveas(gcf,sprintf('Trial%d_CWT_LinearX.png',m));

end
```
